# Blood Pressure and Risk of Atrial Fibrillation: A Mendelian Randomization Study

**DOI:** 10.1101/2020.07.26.20162339

**Authors:** Matthew C. Hyman, Michael G. Levin, Dipender Gill, Venexia M. Walker, Marios K. Georgakis, Neil Davies, Francis E. Marchlinski, Scott M. Damrauer

## Abstract

**Importance:** Observational studies have shown an association between hypertension and atrial fibrillation (AF). Aggressive blood pressure management in patients with known AF reduces overall arrhythmia burden, but it remains unclear whether hypertension is causative for AF.

**Objective:** The primary objective of this study was to investigate the relationship between blood pressure and risk of AF using genetic proxies for blood pressure within a Mendelian randomization (MR) framework. We secondarily explored the relationship between genetically proxied use of anti-hypertensive drugs and risk of AF.

**Design:** Two-sample MR was performed using an inverse-variance weighted meta-analysis with weighted median MR and Egger intercept tests performed as sensitivity analyses. Genetic proxies for the anti-hypertensive drug classes were used to investigate the impact of these therapies on the risk of AF.

**Setting:** International Consortium of Blood Pressure, UK Biobank and Atrial Fibrillation Genetics Consortium.

**Participants:** Summary statistics for systolic blood pressure (SBP), diastolic blood pressure (DBP), and pulse pressure (PP) were obtained from the International Consortium of Blood Pressure and the UK Biobank discovery analysis (>750,000 individuals of European ancestry).

Summary statistics for AF were obtained from the 2018 Atrial Fibrillation Genetics Consortium multi-ethnic GWAS (>65,000 AF cases and >522,000 referents).

**Exposure:** Genetically predicted SBP, DBP and PP as quantified by risk scores.

**Main Outcome:** Odds ratio for AF per 10 mmHg increase in genetically proxied blood pressure.

**Results:** Ten mmHg increases in genetically proxied SBP, DBP or PP were associated with increased odds of AF (SBP: OR 1.17, 95% CI 1.11-1.22, p=1⨯ 10^−11^; DBP: OR 1.25, 95% CI 1.16-1.35, p=3⨯ 10^−8^; PP: OR 1.1, 95% CI 1.0-1.2, p=0.05). Ten mmHg decreases in SBP estimated by genetic proxies of anti-hypertensive medications showed calcium channel blockers (OR 0.66, 95% CI 0.57-0.76, p=8⨯ 10^−9^) and beta-blockers (OR 0.61, 95% CI 0.46-0.81, p=6⨯ 10^−4^) decreased the risk of AF.

**Conclusions and Relevance:** Blood pressure-increasing genetic variants were associated with increased risk of AF, consistent with a causal relationship between blood pressure and AF. These data support the concept that blood pressure reduction through pharmacologic intervention, and specifically calcium channel blockade or beta blockade could reduce the risk of AF.

## Introduction

Atrial fibrillation (AF) remains a leading contributor to cardiovascular morbidity and mortality worldwide.^1,2^ Observational studies have demonstrated an association between modifiable risk factors – specifically hypertension, obesity, alcohol consumption and obstructive sleep apnea – and a reduction in the arrhythmic burden of patients with known AF.^3,4^ Although linked observationally, it is unclear if modification of these risk factors may prevent new onset AF.

Due to its high prevalence, hypertension is thought to be the single greatest contributor to the burden of AF. In population studies such as the Framingham Heart Study and Atherosclerosis Risk in Communities, up to 20% of AF cases are attributed to pre-existing hypertension.^5,6^ Furthermore, 60-80% of patients with known AF have comorbid hypertension.^7^ Despite these observations, initiation of blood pressure lowering therapy was not associated with a clear reduction in AF burden in the Framingham cohort.^8^ Similarly, a randomized comparison of the angiotensin converting enzyme inhibitor, ramipril, versus placebo failed to demonstrate a relationship between ramipril therapy and incident AF.^9^ Secondary analyses in other studies comparing hypertensive agents (angiotensin converting enzyme inhibitors, beta-blockers, calcium channel blockers and diuretics) have not demonstrated a consistent benefit of one anti-hypertensive regimen over another for reducing AF.^10-12^ Comparisons of intensive blood pressure lowering with standard blood pressure lowering have suggested a benefit for patients with hypertension and elevated risk of cardiovascular events, but not hypertension and diabetes.^13,14^ The inconsistent findings of anti-hypertensive therapy studies and observational studies has led some to question the strength of the direct relationship between blood pressure and AF or argue that it is driven by isolated subpopulations.^15-18^

Preventative studies on a population scale are difficult to accomplish in a randomized and adequately powered fashion with sufficient duration. To overcome this limitation, this study employed a population genetics-based approach within a Mendelian randomization framework to better understand the causal role of blood pressure on risk of AF. This technique takes advantage of the random allocation of blood pressure-associated genetic variants that occurs at conception. This random assortment minimizes the chance of environmental confounding, enabling investigation into the causal relationship between blood pressure and AF. We subsequently evaluated genetic proxies for the pathways targeted by anti-hypertensive medications to better understand potential class effects of anti-hypertensive medications on AF.

## Methods

### Study populations

For the primary analysis, summary-level data for genome wide association studies (GWAS) of hypertension and AF were used.^19,20^ Blood pressure data were obtained from the 2018 Evangelou et al. International Consortium for Blood Pressure + UK Biobank GWAS meta-analysis, which included systolic blood pressure, diastolic blood pressure, and pulse pressure measurements in up to 757,601 individuals. Summary statistics for blood pressure are publicly available, and were downloaded from the NHLBI GRASP catalog (https://grasp.nhlbi.nih.gov/FullResults.aspx). AF data was obtained from the 2018 Roselli et al. atrial fibrillation GWAS meta-analysis from the Atrial Fibrillation Genetics (AFGen) consortium study, including 65,446 AF cases and 522,744 controls. Summary statistics for AF were contributed by the AFGen consortium (http://afgen.org), are publicly available, and may be downloaded from the Variant to Function Knowledge Portal (http://www.kp4cd.org/datasets/v2f). Because both the blood pressure exposure and AF outcome studies included participants from UK Biobank (458,577 for BP and 351,017 for AF), bias due to sample overlap was estimated.^21^

### Study exposures

The 2018 Evangelou et al. International Consortium for Blood Pressure + UK Biobank discovery meta-analysis GWAS included up to 757,601 participants. This analysis included up to 299,024 European participants from 77 independent studies genotyped with various arrays and imputed to either the 1000 Genomes Reference Panel or the HRC platforms, and 458,577 participants from the UK Biobank. Blood pressure ascertainment varied among cohorts, and study-specific details are presented in the supplemental material.^19^ For each BP trait, genetic variants associated with systolic blood pressure (SBP), diastolic blood pressure (DBP), and pulse pressure (PP) at genome-wide significance (p < 5×10^−8^) were identified and LD-pruned (distance threshold = 10,000kb, r^2^ < 0.001) using the 1000 Genomes European-ancestry reference panel to identify independent variants. Because the Evangelou et al. study adjusted effect estimates for body mass index potentially leading to introduction of collider-bias as body mass index is causal for both elevated blood pressure and AF, a sensitivity analysis was performed using systolic (N = 436,419) and diastolic (N = 436,424) blood pressure genome-wide association study summary statistics from European UK Biobank participants adjusted for genotyping array, sex, and population structure.^22^

### Primary outcome

The AFGen consortium identified participants from more than 50 studies (84.2% European, 12.5% Japanese, 2% African American, and 1.3% Brazilian and Hispanic), including participants from the UK Biobank, Biobank Japan, other international biobanks, and international cardiovascular cohort studies (adjusted for age, sex, and study-specific covariates). AF ascertainment was study-specific, including diagnostic codes, electronic health record information and self-report.

### Study design

The primary analysis estimated the effect of blood pressure on risk of AF using two-sample Mendelian randomization with an inverse-variance weighted model with random effects. The MR-Egger bias intercept test was used to identify the presence of bias from directional pleiotropy. Sensitivity analysis was performed using weighted median MR and Egger intercept tests, which are more robust to the presence of invalid genetic instruments.^23^

Recent work has demonstrated that genetic proxies can be used to estimate the effect of individual anti-hypertensive drug classes on clinical outcomes using a Mendelian randomization framework.^24,25^ We used two approaches to estimate the effect of blood pressure lowering medication on risk of AF:

1. Genes encoding the targets of anti-hypertensive medications (angiotensin-converting enzyme inhibitors, angiotensin receptor blockers, β-blockers, calcium channel blockers and thiazide diuretic agents) were identified using DrugBank and the GeneHancer database in the GeneCards platform (v4.7).^24^ SNPs were identified within corresponding genes, promoter regions, or enhancers that were associated with SBP at genome-wide significance (P<5×10^−8^) and clumped to a linkage disequilibrium (LD) threshold of r^2^<0.1 using the 1000G European reference panel. These genetic variants were used as instruments to model the effect of lower SBP mediated by individual anti-hypertensive drug classes. The SNPs were then utilized to estimate the effect of the individual anti-hypertensive drug classes on risk of AF using two-sample inverse variance weighted and median weighted Mendelian randomization as above.
2. Expression quantitative trait loci (eQTL) for protein targets of antihypertensive medications were used as a proxy for the action of a drug on its target (*e*.*g*. variants associated with angiotensin-converting enzyme gene expression as a proxy for the angiotensin converting enzyme inhibitor drug class).^25^ Twelve antihypertensive drug classes were considered: adrenergic neuron blocking drugs; alpha-adrenoceptor blockers, angiotensin-converting enzyme inhibitors, angiotensin-II receptor blockers, beta-adrenoceptor blockers, calcium channel blockers, centrally acting antihypertensive drugs, loop diuretics; potassium-sparing diuretics and aldosterone antagonists, renin inhibitors, thiazides and related diuretics and vasodilator antihypertensives. SNPs were identified for the protein targets of each drug class using the GTEx project data (Release V7; dbGaP Accession phs000424.v7.p2), which contains expression quantitative trait loci analyses of 48 tissues in 620 donors.^26^ SNPs defined by GTEx as the variant with the smallest nominal p-value for a variant-gene pair were selected for analysis and validated as instruments by estimating their effect on systolic blood pressure using two-sample Mendelian randomization. SNPs with evidence of an effect on systolic blood pressure were used for the analysis.

### Statistical Analysis

Two-sample Mendelian randomization was performed using the TwoSampleMR package in R (https://github.com/MRCIEU/TwoSampleMR).^27^ Variants associated with each blood pressure exposure at genome-wide significance (p < 5×10-8) were harmonized with the variants from the atrial fibrillation GWAS^20^, and LD-clumped (distance threshold = 10,000kb, r^2^ = 0.001) using the 1000 Genomes European ancestry reference panel, identifying a final set of independent single-nucleotide polymorphisms (SNPs) to use as a genetic instrument for blood pressure. Inverse variance weighted two-sample Mendelian randomization with random effects was used as the primary analysis with a weighted median analysis performed as a sensitivity analyses.^28^ For each variant included in the genetic instruments, the proportion of variance (R^2^) in the phenotype explained was calculated using the formula 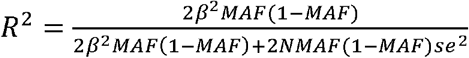 (where MAF represents the effect allele-frequency, beta represents the effect estimate of the genetic variant in the exposure GWAS, *se* represents the standard error of effect size for the genetic variant, and N represents the sample size).^29^ F-statistics were then calculated for each variant using the formula 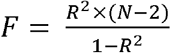 to assess the strength of the selected instruments.^30^ All statistical analyses were performed using R version 3.6.2.31.

## Results

### Association of blood pressure with atrial fibrillation

We identified a set of independent variants to serve as instruments for systolic blood pressure (n=399) and diastolic blood pressure (n=398), and pulse pressure (n=347) which accounted for 4.0%, 4.2% and 3.6% of the measured variability in these exposures, respectively (**eTable 1-3**). For the systolic blood pressure instrument, the mean F-statistic was 75 (range 30.4-645.7). For the diastolic blood pressure instrument, the mean F-statistic was 79.9 (range 30-846.6). For the pulse pressure instrument, the mean F-statistic was 76.4 (range 30.4-627.9). Bias due to sample overlap from UK Biobank participants included in both the blood pressure exposure GWAS and AF outcome GWAS was estimated to be negligible across a range of observational effect sizes: for example, at 100% sample overlap, bias was estimated to be 0.0003 for an observational odds ratio of 1.3 and 0.00069 for an observational odds ratio of 1.6 (**eTable 4**).

Two-sample Mendelian randomization using the above genetic instruments and inverse variance-weighted modeling demonstrated that each 10mmHg genetically predicted increase in systolic blood pressure, diastolic blood pressure and pulse pressure increased the risk of AF (SBP: OR 1.17, 95% CI 1.11-1.22, p=1⨯ 10^−11^; DBP: OR 1.25, 95% CI 1.16-1.35, p=3⨯ 10^−8^; PP: OR 1.1, 95% CI 1.0-1.2, p=0.05) (**Figure 1**). Results were similar in a sensitivity analysis using the weighted median method, with increased SBP, DBP and PP increasing risk of AF (SBP: OR 1.18, 95% CI 1.12-1.23, p=5⨯ 10^−11^; DBP: OR 1.24, 95% CI 1.14-1.34, p=4⨯ 10^−7^; PP: OR 1.11, 95% CI 1.02-1.2, p=0.01) (**Figure 1**). The effects of SBP and DBP on risk of AF were also similar using alternative genetic instruments derived from UK Biobank (**eFigure 1**).

**Figure 1:**
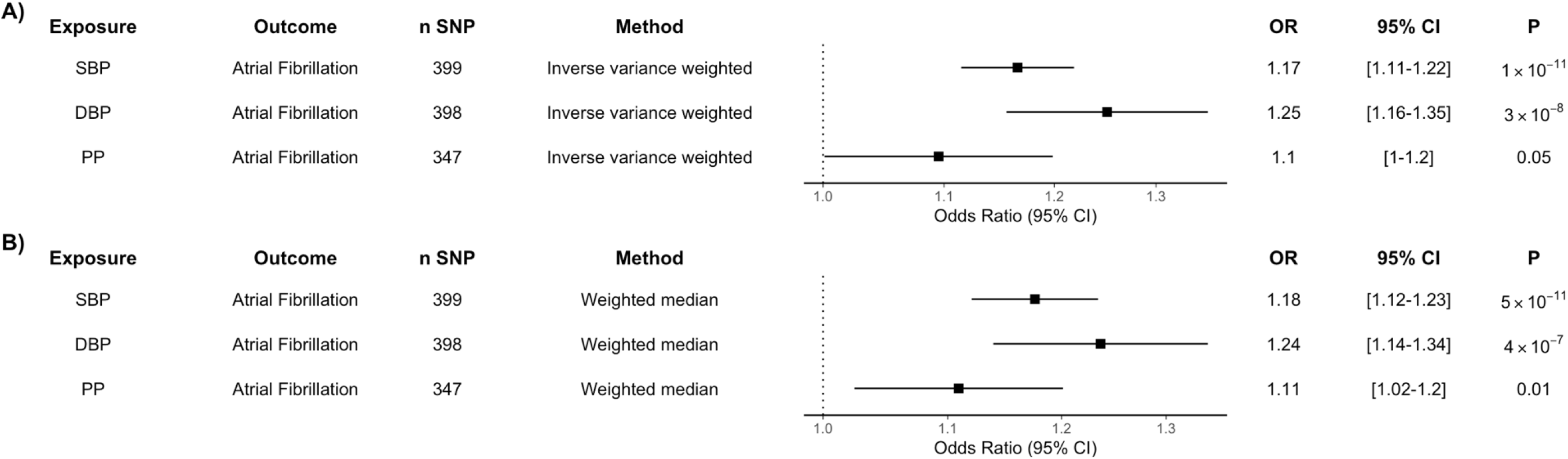
Genetic Proxies of Blood Pressure and Risk of Atrial Fibrillation. A) Two-sample Mendelian randomization using an inverse variance weighted model was created using a genetic instrument associated with a 10 mmHg increase in systolic blood pressure (SBP), diastolic blood pressure (DBP) and pulse pressure (PP) and risk of atrial fibrillation. B) A median weighted model was created as a sensitivity analysis. Figures are expressed as Odds Ratios (OR), 95% Confidence Intervals (CI) and P-values for Mendelian randomization estimates.

**eFigure 1:**
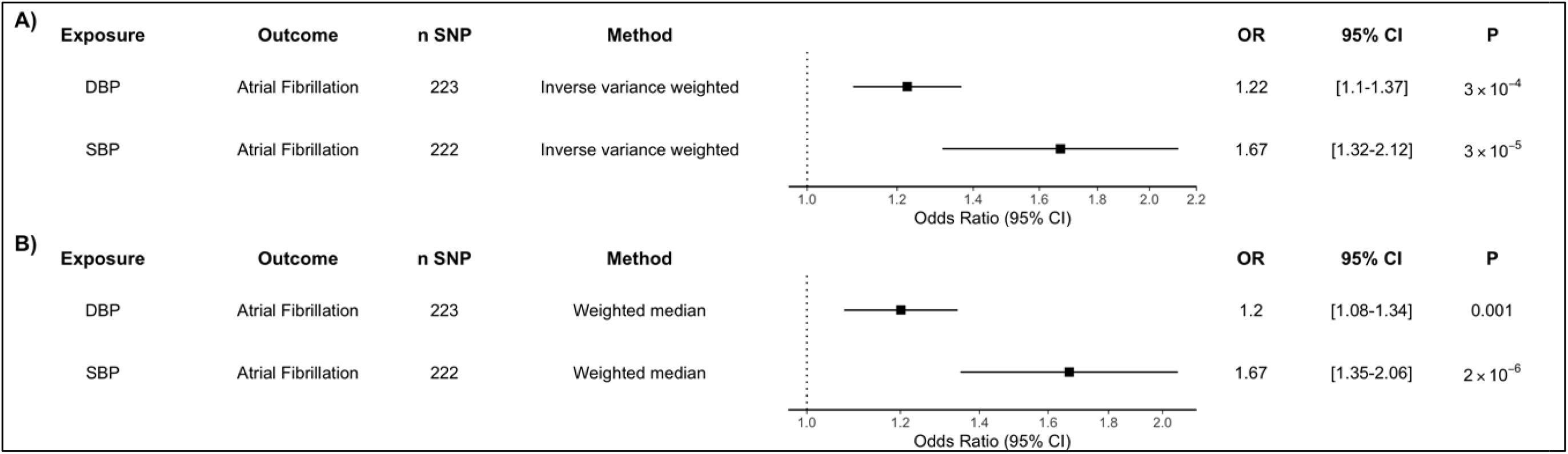
Effect of Systolic and Diastolic Blood Pressure on AF Risk using UK Biobank Blood Pressure Genetic Instruments. A) Two-sample Mendelian randomization using an inverse variance weighted model was created using a genetic instrument associated with a 10 mmHg increase in systolic blood pressure (SBP) or diastolic blood pressure (DBP) and risk of atrial fibrillation. B) A median weighted model was created as a sensitivity analysis. Figures are expressed as Odds Ratios (OR), 95% Confidence Intervals (CI) and P-values for Mendelian randomization estimates.

### Genetically proxied blood pressure reduction through antihypertensive drug targets and atrial fibrillation

To estimate the effect of blood pressure reduction by different classes of anti-hypertensive medications, we identified common genetic variants located within genes of protein targets of calcium channel blockers and beta blockers, as previously described.^24^ Twenty independent variants within protein targets of calcium channel blockers (CCBs) and 5 independent variants within protein targets of beta blockers (BBs) were associated with systolic blood pressure at genome-wide significance **(eTable 5)**. When using these genetic proxies to estimate the effect of each 10 mmHg decrease in systolic blood pressure by each anti-hypertensive drug class, genetically predicted protein targets of CCBs and BBs were associated with lower risk of AF (CCB: OR 0.66, 95% CI 0.57-0.76, p=8×10-9; BB: OR 0.61, 95% CI 0.46-0.81, p=6×10-4; **Figure 2**). In a complimentary analysis, we employed eQTLs for the protein targets of anti-hypertensive medications given genetic variants may exert their action via distant interactions (rather than via a true cis-acting association) **(eTable 6)**.^25^ Using this technique, anti-hypertensive medication proxies reduced the risk of AF (CCB: SNP *n* = 23, OR 0.66, 95% CI 0.57 – 0.76, p = 8⨯ 10^−9^; BB: SNP *n* =10, OR 0.61, 95% CI 0.46 – 0.81, p = 6⨯ 10^−4^). There was not strong evidence of effect of other anti-hypertensive medication classes on AF risk (**eTable 7)**.

**Figure 2:**
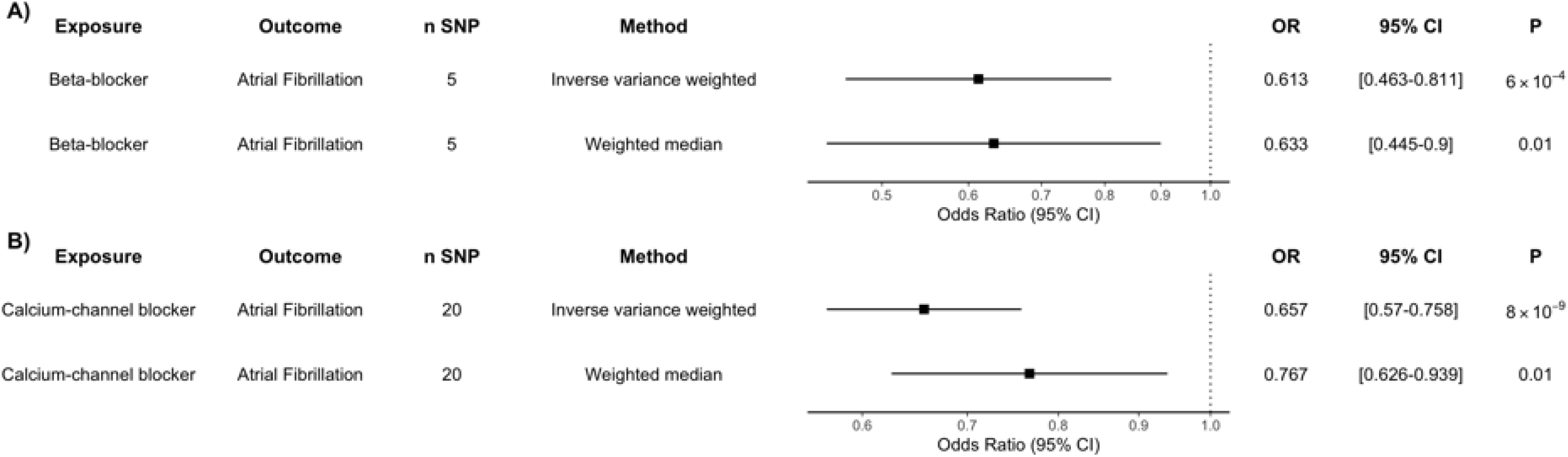
Genetic Proxies of Anti-Hypertensive Medications and Atrial Fibrillation Risk. Two-sample Mendelian randomization was performed using genetic proxies for 10 mmHg systolic blood pressure lowering by individual anti-hypertensive medication classes: A) beta-blockers and B) calcium channel blockers. Inverse variance weighted and median weighted models are presented. Figures are expressed as Odds Ratios (OR), 95% Confidence Intervals (CI) and P-values for Mendelian randomization estimates.

## Discussion

This study utilized Mendelian randomization to leverage population-level genetic information to explore the causal relationship between blood pressure and AF. The genetic determinants of elevated systolic blood pressure, diastolic blood pressure and pulse pressure were found to strongly associate with risk of AF; an association that persisted in statistical sensitivity analyses more robust to the inclusion of pleiotropic variants. Previously validated genetic proxies for the therapeutic effects of anti-hypertensive drug classes were used to estimate the impact of individual anti-hypertensive drug classes on incident AF, suggesting a potential role for anti-hypertensive medications in prevention.

A relationship between hypertension and AF has previously been established in observational analyses.^5,6^ These findings, however, were limited in demonstrating a causal role for hypertension in the development of AF due to the potential of residual confounding and reverse causation.^15^ This study sought to mitigate this risk of confounding by using genetic instruments randomly assorted in the population to proxy the effect of increased blood pressure traits on risk of AF. In doing so, we found that across all the considered measures of blood pressure (systolic blood pressure, diastolic blood pressure and pulse pressure), higher genetically proxied levels were associated with increased the risk of AF. These elevated blood pressure effects are directionally similar to those identified in observational studies and previous analyses using polygenic risk scores in smaller cohorts of patients further lending strength to the causal relationship between blood pressure and AF.^31-33^

A variety of mechanisms have been proposed to explain how hypertension contributes to risk of AF. Animal models of hypertension have demonstrated the presence of left atrial scaring and inflammation.^34-36^ This scaring and fibrosis is thought to create altered patterns of conduction and functional slowing allowing for the development and perpetuation of AF triggers.^35,37,38^ Concordantly, hypertensive animals have greater heterogeneity of atrial activation with increased susceptibility to AF induction.^35^ Other manifestations of left atrial remodeling such as increased left atrial size have been associated with hypertension and elevated systolic blood pressure in particular.^39^ It should be noted, however, that the impact of hypertension on AF risk persists after adjustment for left atrial size and mass.^31^

Beyond mechanism, this study explores the question of whether pharmacologic intervention may meaningfully impact a patient’s risk of AF. While it would be ethically difficult to fully withhold anti-hypertensive therapy in a randomized trial, we leveraged genetic proxies to explore the impact of individual anti-hypertensive drug classes. Our study suggests that both calcium channel blocker and beta-blockers can significantly mitigate a patient’s risk of developing AF, though neither class was more effective than the other. There are inherent limitations to comparing genetic proxies for a lifetime of blood pressure lowering by an anti-hypertensive drug class to clinical trials that represent a limited duration of anti-hypertensive therapy. With that being said, our findings are consistent with clinical trials and case-control analyses that have not found consistent improvement in AF burden when beta-blockers and calcium channel blockers are compared to other drug classes including angiotensin-receptor blockers and angiotensin-converting enzyme inhibitors.^11,12,40^

### Limitations

First, the GWAS of hypertension and AF were multiethnic, though the underlying studies were enriched with individuals of European ancestry. This may have skewed the risk estimates in our findings, and as such the analysis should be repeated in other populations before being generalized across ethnic groups. Second, this analysis estimates the lifelong effects of genetically predicted blood pressure reduction on AF risk and does not directly investigate effects of shorter-term alterations in blood pressure such as through pharmacological treatment in adulthood. Third, it should be noted that while the risk reduction of calcium channel blockers and beta-blockers was quantified in terms of 10 mmHg blood pressure increments and in doing so assumes a linear model. Consequently, this study cannot answer the question of what level of blood pressure reduction maximizes AF risk reduction.

## Conclusions

Blood pressure-increasing genetic variants were associated with increased risk of AF, consistent with a causal relationship between blood pressure and AF. These data support the concept that blood pressure reduction through pharmacologic intervention, and specifically calcium channel blockade or beta blockade could reduce the risk of AF.

## Data Availability

All data included in this study were obtained from publicly available sources.

https://grasp.nhlbi.nih.gov/FullResults.aspx

http://www.kp4cd.org/datasets/v2f

## Author Contributions

Drs Hyman and Levin contributed equally. Dr. Levin had full access to all of the data in the study and takes responsibility for the integrity of the data and the accuracy of the data analysis.

## Study concept and design

Hyman, Levin, and Damrauer.

## Acquisition, analysis, or interpretation of data

Hyman, Levin, Gill, Walker, Georgakis, Davies and Damrauer.

## Drafting of the manuscript

Hyman, Levin and Damrauer.

## Critical revision of the manuscript for important intellectual content

Hyman, Levin, Gill, Walker, Georgakis, Davies, Marchlinski, and Damrauer.

## Statistical analysis

Levin.

## Abbreviations

AF: atrial fibrillation
BB: beta-blocker
CCB: calcium channel blocker
DBP: diastolic blood pressure
eQTL: Expression quantitative trait loci
LD: linkage disequilibrium
MR: Mendelian randomization
PP: Pulse pressure
SBP: systolic blood pressure

